# Molecular models of multiple sclerosis severity identify heterogeneity of pathogenic mechanisms

**DOI:** 10.1101/2020.05.18.20105932

**Authors:** Christopher Barbour, Peter Kosa, Mihael Varosanec, Mark Greenwood, Bibiana Bielekova

**Affiliations:** Neuroimmunological Diseases Section, National Institute of Allergy and Infectious Diseases, National Institutes of Health, Bethesda, MD; Department of Mathematical Sciences, Montana State University, Bozeman, MT

## Abstract

The inability to measure putative pathogenic processes in the central nervous system (CNS) of living subjects precludes the determination of their temporal distribution, intra-individual heterogeneity, and their ability to predict disease course. Using multiple sclerosis (MS) as an example of a complex neurological disorder, we sought to determine if cerebrospinal fluid (CSF) biomarkers can be aggregated to predict future rates of MS progression and provide molecular insight into mechanisms of CNS destruction. 1,305 CSF biomarkers were analyzed blindly in the longitudinal training dataset (N=129) of untreated MS patients, using DNA-aptamer assay. Random forest models, validated in an independent longitudinal cohort (N=64), uncovered signatures of MS severity, measured by clinical scales and volumetric brain imaging. Cluster analysis revealed intra-individual molecular heterogeneity of disease mechanisms that include both CNS- and immune-related pathways and may represent novel targets for inhibiting MS progression.

## Introduction

Successful management of chronic, polygenic diseases requires patient-specific polypharmacy regimens that target all pathogenic mechanisms underlying disease expression in the patient. This strategy is feasible in e.g., cardiovascular diseases, where the contributing pathogenic mechanisms are easily measured. In contrast, it is currently impossible to measure diverse pathogenic mechanisms contributing to the destruction of the central nervous system (CNS). This limits new drug development and makes clinical management of patients suboptimal. Advances in proteomics allow for accurate measurements of thousands of proteins in cerebrospinal fluid (CSF) (1, 2). Using DNA-aptamer-based technology (i.e., SOMAscan©(3); Somalogic Inc, Boulder, CO, USA) combined with machine learning, we have published a molecular diagnostic test of multiple sclerosis (MS) (4) that greatly outperforms magnetic resonance imaging (MRI)-based diagnosis of MS (i.e., independent cohort-validated area under receiver-operator characteristic curve (AUROC) 0.98 for the molecular diagnostic test (4) versus AUROC of ~0.70 for the MRI-based tests (5)). In recognition of the insufficient accuracy of MRI-based diagnosis, the 2017 revision of MS diagnostic criteria incorporates a possibility to evaluate CSF oligoclonal bands (OCB) (6). This opens an opportunity to bring to clinical practice advanced laboratory tests, such as SOMAscan, which provides diagnostic advantage and may pin-point patient-specific pathophysiological drivers of disability.

Pathologists identified multiple processes in MS CNS tissue autopsy, but it remains unclear which of these are pathogenic versus representan epiphenomenon. Intrathecally-compartmentalized inflammation (7), associated with the tertiary lymphoid follicles, has strong evidence for pathogenicity, based on correlations with rates of disability progression (8), although this observation lacks independent validation. CNS compartmentalized inflammation might be identified in approximately 30% of subjects with MS (irrespective of their categorization into relapsing-remitting [RRMS], secondary-[SPMS] or primary-progressive MS [PPMS]), as meningeal enhancement using post-contrast fluid-attenuated inversion recovery (FLAIR) MRI (9). The accuracy of this imaging biomarker is limited by observation of meningeal enhancement in healthy volunteers (HVs). CNS compartmentalized inflammation can be measured using a research-based CSF-biomarker test (7), which demonstrated high prevalence in people with progressive MS and no positive values in HVs (7). Nevertheless, this cumbersome assay is not translatable to broad clinical practice.

Non-immune mechanisms such as mitochondrial dysfunction, hypoxia, oxidative stress, demyelination, toxic (A1) astroglial activation (10,11), and neuronal death might also be measured by CSF biomarkers. The most promising of these is neurofilament light chain (NFL) (12), detectable in HVs, but in greater quantities in neurodegenerative diseases. NFL correlates strongly with MS relapses or contrast enhancing lesions (CELs) but has weak predictive power for future disability progression. Thus, there remains a need for development of biomarkers reflective of diverse molecular intrathecal processes with proven clinical value.

This paper tests the hypothesis that CSF biomarker models provide insight into MS pathophysiology, identify molecular disease heterogeneity, and lead to an independent-cohort validated prognostic test.

## Results

### Adjusting Somamers based on physiological age and gender associations

Because CNS processes that may participate in disability progression (e.g., compartmentalized inflammation, demyelination, axonal loss) evolve with MS duration and patients’ age, simple statistical adjustments for age may limit detection of MS-specific relationships. Similarly, because females are overrepresented in RRMS in comparison to PPMS, statistical adjustments for gender may also mask identification of processes contributing to MS severity. Instead, we sought to differentiate the natural aging process and physiological gender differences from MS-specific processes.

We merged the analysis of HVs in this study with a published cohort of 3301 HVs from the INTERVAL study analyzing serum using SOMAscan (1) to identify age- and gender-related Somamers. Thus, we examined the age- and gender-related Somamers from INTERVAL cohort for: 1. Concordant directionality in the relationships (p-value < 0.05) between INTERVAL HV and our HV cohort; and 2. Statistically significant relationship with age and/or gender in our MS cohort. Using these two criteria, 73 age-associated Somamers were identified. Considering that aging may affect CNS proteins that may not be accurately measured in serum, we identified two additional Somamers in our HV CSF cohort with strong evidence of age associations after Bonferroni adjustments (PGF and SLPI), that were not identified in the INTERVAL study. Out of these 75 HV-age-associated biomarkers, 22 (29.3%) showed discordant changes between the two HV cohorts and the MS cohort (i.e., increasing with age in MS patients and decreasing with age in HVs) (Figure 1A).

**Figure 1:**
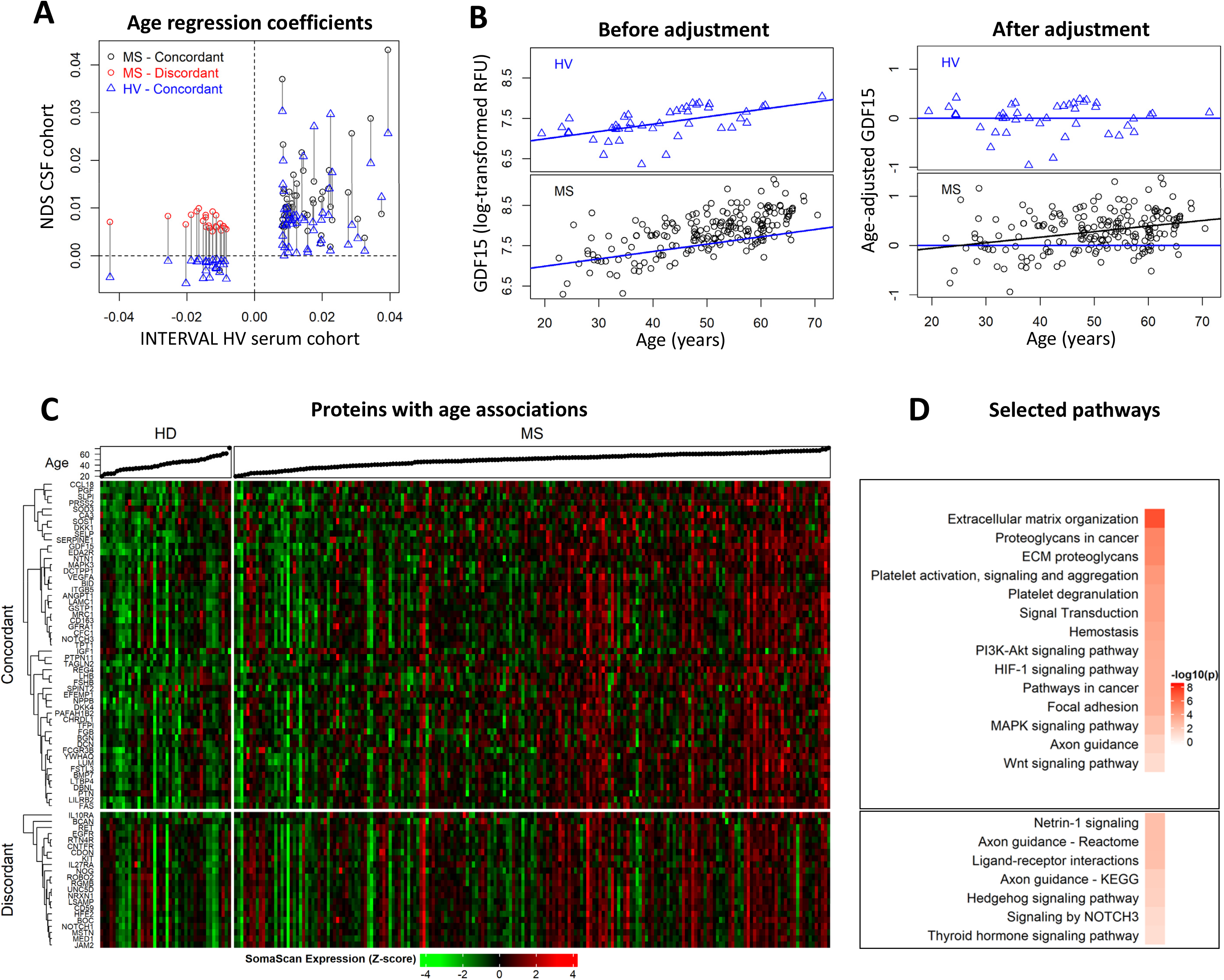
Adjusting Somamers based on physiological age associations. **A)** Regression coefficients for the 75 Somamers with age associations from cerebrospinal fluid (CSF) and serum. The x-axis represents the coefficients for age from the healthy volunteer (HV) serum in the INTERVAL cohort and the y-axis represents the coefficients from our cerebrospinal fluid cohort. Vertical lines connect the CSF coefficients for our multiple sclerosis (MS) and HV cohorts for the same Somamer. Black circles correspond to MS CSF coefficients with concordant associations with age compared to HV, red circles correspond to MS CSF coefficients with discordant associations with age compared to HV, and blue triangles correspond to HV CSF coefficients with concordant associations with age compared to HV CSF. **B)** Demonstration of age adjustments to MS CSF values on an example of log-scaled values for GDF15. GDF15 values (y-axis) versus age (x-axis) are displayed for both HVs (top) and MS (bottom) cohorts, before (left) and after (right) adjustment. The HV simple linear regression line is superimposed on each panel as a blue line, which was used to perform the adjustments. The black line in the bottom-right panel corresponds to the regression line of adjusted GDF15 versus age in MS patients, indicating that after adjustment, GDF15 has an additional age association in MS patients. **C)** Heatmap displaying the standardized expression (log-scaled z-scores) for the 75 selected Somamers (rows), separated based on HV/MS concordance or discordance, for all patient samples (columns). Corresponding ages for each participant are displayed in ascending order at the top of the heatmap. **D)** Selected pathways identified using STRING analysis along with FDR-adjusted −loglO(p-values) for age concordant and discordant proteins, respectively. See also Table S2 and Table S3.

To verify that identified proteins are indeed age-related based on current knowledge, we used the Search Tool for the Retrieval of Interacting Genes/Proteins (STRING) annotations. Reassuringly, STRING of concordant age-related proteins (Figure 1 and Table S2) identified Kyoto Encyclopedia of Genes and Genomes (KEGG) and Reactome pathways previously associated with physiological aging, such as proteoglycans/chondroitin sulfate and extracellular matrix reorganization, signaling pathways p53, PI3K-AKT, MAPK, HIF-1 and WNT, apoptosis and Alzheimer’s disease. While most of the age-concordant proteins were proteins secreted to extracellular space and were part of the extracellular matrix, age-discordant CSF proteins (i.e., decreased in HV but increased in MS) belonged to two categories: 1. Secreted proteins linked to immune system and 2. the cell surface/membrane-anchored proteins found in axons and the neuronal cell body (Figure 1 and Table S3), suggesting destruction of these structures in MS. The pathways enriched for age-discordant CNS proteins are metabolism, axon guidance, netrin-1, NOTCH, hedgehog, and thyroid hormone signaling, all linked to neurogenesis or myelination. 35 Somamers were detected to have physiological gender differences, with all but one (SERPINA10) showing concordant differences between MS patients and HVs (Figure 2). STRING analysis again confirmed validity of our approach: the 7 proteins elevated in women are related to ovulation, ovarian steroidogenesis, and prolactin signaling (Figure 2 and Table S4). In contrast, male-elevated proteins are linked to immunity (innate immunity, chemokines), fluid shear stress, and atherosclerosis (Figure 2 and Table S5).

**Figure 2:**
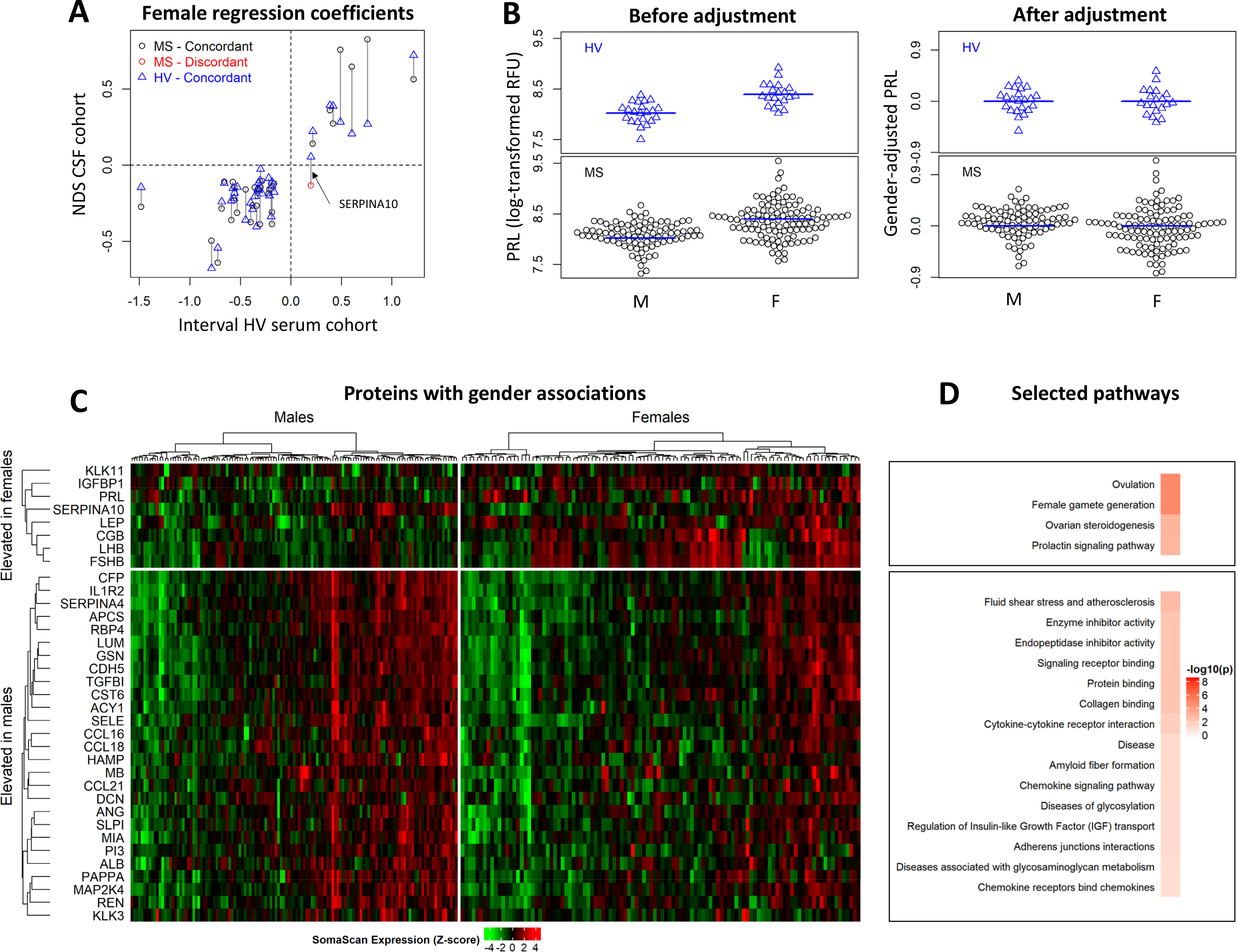
Adjusting Somamers based on physiological gender associations. **A)** Regression coefficients for the 35 Somamers with gender associations from cerebrospinal fluid (CSF) and serum. The x-axis represents the coefficients for female from the healthy volunteer (HV) serum in the INTERVAL cohort and the y-axis represents the coefficients from our cerebrospinal fluid cohort. Vertical lines connect the CSF coefficients for our multiple sclerosis (MS) and HV cohorts for the same Somamer. Black circles correspond to MS CSF coefficients with concordant associations with gender compared to HV, red circles correspond to MS CSF coefficients with discordant associations with gender compared to HV, and blue triangles correspond to HV CSF coefficients with concordant associations with gender compared to HV CSF. Only a single Somamer, SERPINA10 identified by a black-arrow, showed concordance differences between HVs and MS patients, and was excluded from consideration. **B)** Demonstration of gender adjustments to MS CSF values on an example of log-scaled values for PRL. PRL values (y-axis) versus gender (x-axis) are displayed for both HVs (top) and MS (bottom) cohorts, before (left) and after (right) adjustment. The HV averages is superimposed on each panel, which was used to perform the adjustments. **C)** Heatmap displaying the standardized expression (log-scaled z-scores) for the 35 selected Somamers (rows), separated based on elevation in females/males, for all patient samples, separated by males and females (columns). **D)** Selected pathways identified using STRING analysis along with FDR-adjusted −loglO(p-values) for female elevated and male elevated proteins, respectively. See also Table S4 and Table S5.

To account for the physiological aging process and sexual dimorphism in our analyses, we adjusted all age- and gender-related Somamers using linear regression models derived from our HV cohort as described in STAR Methods and used these age- and gender-adjusted values for all further analyses.

### MS is not associated with accelerated aging

Among the proposed hypotheses of MS progression is the idea that MS patients suffer from accelerated aging (13). Thus, we tested the hypothesis that the CSF proteomic signature of physiological aging estimates higher than biological age for MS patients. To this end we used a regularized multiple linear regression model to develop a CSF biomarker-based model of chronological age in HV cohort (Figure 3A). When applied to the MS cohort and examining phenotypical subtypes of MS separately, we observed that the HV based model of physiological aging tends to overestimate age in RRMS (without reaching statistical significance) and, surprisingly, underestimate physiological age in both progressive MS subtypes (Figure 3B). Thus, we conclude that molecular mechanisms different from (accelerated) physiological aging are responsible for CNS tissue loss in MS.

**Figure 3:**
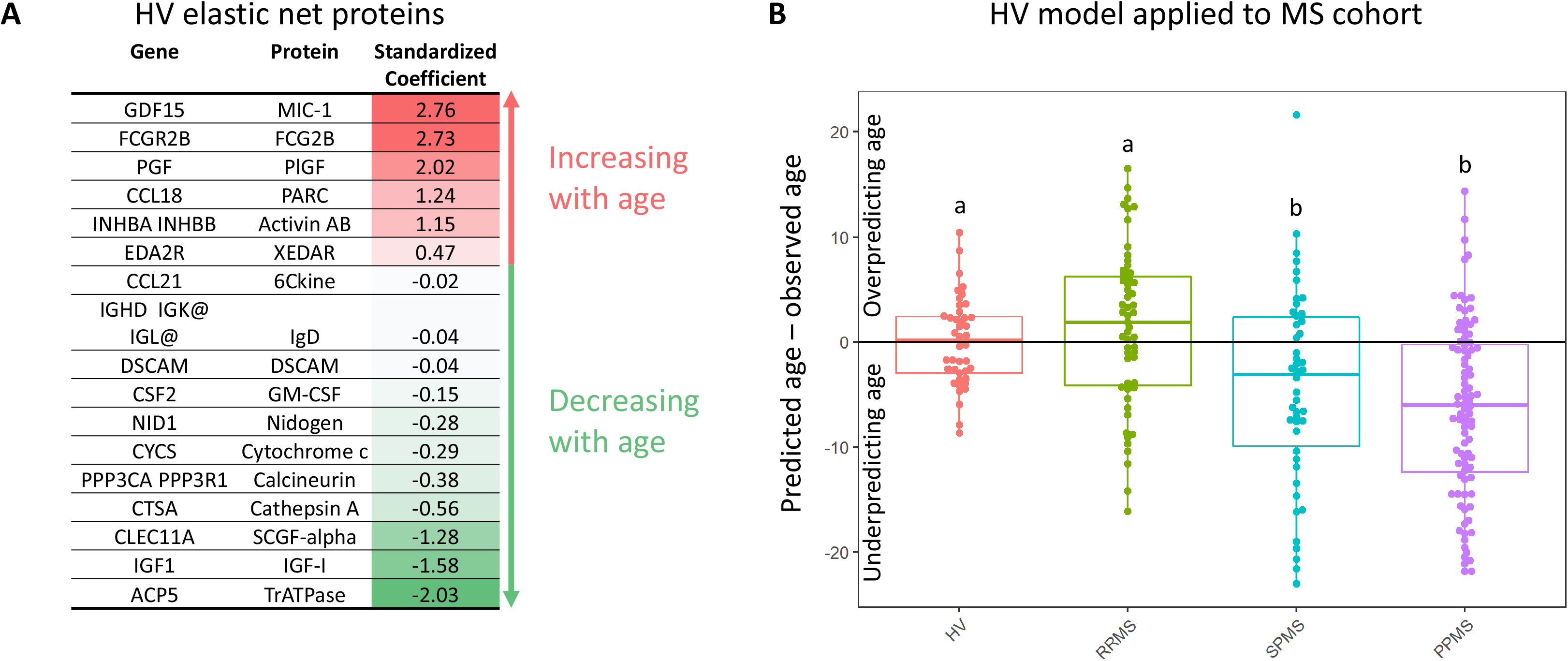
MS is not associated with accelerated aging. **A)** Standardized regression coefficients from developing an elastic net model predicting age using all available Somamers in the healthy volunteer (HV) cohort. Red-shading corresponds to Somamers estimated to increase with age, and green-shading corresponds to Somamers estimated to decrease with age. **B)** Difference between model predicted ages and observed ages (y-axis) based on HV data across multiple sclerosis (MS) subtypes (x-axis). A compact letter display is used to display the results of all pairwise comparisons using Tukey’s honest significant difference. When this model is applied to the MS cohort, the age tends to be slightly overpredicted in RRMS compared to HV (not statistically significant) and underpredicted in the progressive MS subtypes compared to HV and RRMS (p-value <0.05).

### Identifying molecular pathways associated with MS severity

Biological processes that correlate with MS disability may be a consequence, rather than driver of CNS destruction. To facilitate identification of causative processes, we aimed to identify pathways that correlate with MS progression rates, measured by MS severity outcomes. Unfortunately, EDSS-based MS severity outcomes MS Severity Score (MSSS (14)) and Age Related MS Severity Score (ARMSS (15)) have weak power to predict future rates of disability progression (16). Therefore, we used a more accurate MS Disease Severity Score (MS-DSS (16)), calculated at the first untreated clinic visit (concomitantly with CSF collection) as the primary outcome against which we modelled CSF biomarkers. This allowed us to test predictive power of CSF-biomarker-based models against future clinical progression slopes in a de facto prospective manner (Figure 4A). As sensitivity analyses for the robustness of the gained biological insight, we included two secondary outcomes: 1. MS-DSS collected at the last clinical follow-up, where we utilized automated computation of disability scores using the NeurEx App (17). By eliminating scoring differences among clinicians, NeurEx-based outcomes are more accurate. 2. MRI-based MS severity outcome calculated as residuals from the linear regression model of brain atrophy (i.e., 1- brain parenchymal fraction (BPFr); Figure 4B) against age, performed in the untreated stage within 3 months of CSF collection. Patients with higher MRI severity have lost brain tissue at a faster rate than their equally-aged peers.

**Figure 4:**
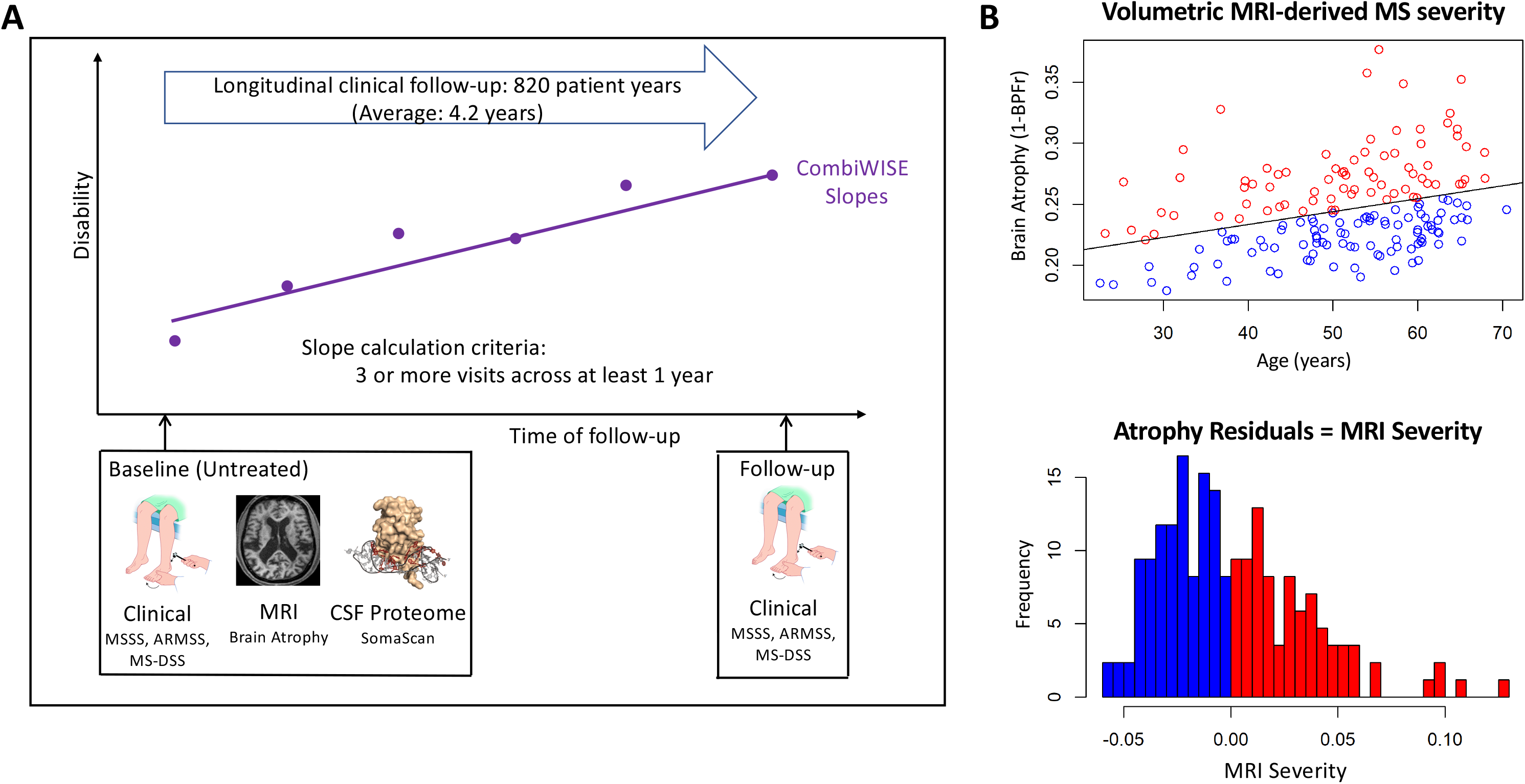
Schematic of clinical data collection and MRI severity calculation. **A)** Visual depiction of the data collection timeline. During the first untreated visit for each patient, clinical, magnetic resonance imaging (MRI), and cerebrospinal fluid (CSF) were collected. Patients were followed for an average of 4.2 years, with disability outcomes being collected. At the final visit, clinical severity outcomes were recorded, and patient-specific disability slopes for CombiWISE were calculated via simple linear regression. **B)** Visual depiction of the calculation of the magnetic resonance imagining (MRI) severity measure. Brain atrophy, measured as 1 - brain parenchymal fraction (BPFr) (y-axis), was regressed against age (y-axis) for baseline visit data (top). MRI severity was calculated at baseline as the residuals from this relationship (bottom). Positive residuals (red) from this regression indicate more brain atrophy than the expected atrophy given a patient’s age and negative residuals (blue) indicate less brain atrophy than the expected atrophy given a patient’s age.

To gain biological insight about processes that correlate with MS severity, we used Gene Set Enrichment Analysis (GSEA) that uses all measurements of CSF biomarkers, not just proteins that correlate with the outcomes in formal statistical analysis. Nevertheless, we provide Spearman correlation coefficients and False Discovery Rate (FDR)-adjusted p-values (18) for individual Somamers in the Table S6. The following number of Somamers had adjusted p-values < 0.05 for correlation with tested outcomes: 55 for MRI MS severity model, 76 for MS-DSS measured at follow-up visit and 26 for MS-DSS measured at baseline visit. Only 2 Somamers correlated with ARMSS at baseline and 1 at follow-up visits and no Somamers correlated with MSSS.

In addition to GSEA analysis based on measured Spearman correlation coefficients, we also provide FDR adjusted p-values (or q-values) from STRING analysis (19) results for positively- and negatively-correlating proteins to MS severity outcomes. These sensitivity analyses provide reassuringly overlapping results in biological interpretations of MS-severity related processes. Due to space limitations we’ll focus on most relevant findings.

For MS severity outcomes, the complement and coagulation cascade (q-values ranging from 5.88 e-9 to 3.50 e-3) and overlapping Staphylococcus infection pathway (q-value = 2.47 e-6) are positively-associated pathways. The negatively-associated processes are axon guidance and neuron differentiation (q-values 3.40 e-3 to 5.20 e-3) and overlapping NOTCH signaling pathway (q-values ranging from 7.80 e-4 to 3.50 e-3). Additional processes that inversely correlated with MS severity are mitochondrial biogenesis (q-value = 5.90 e-3) and respiratory electron transport (q-value = 4.6 e-3).

The excellent reproducibility of biological processes identified across all three outcomes of MS severity was corroborated in our sensitivity analyses of pathway enrichment based solely on proteins with statistically-significant correlations: complement and coagulation cascades were again dominating positive correlations with MS severity: proteins positively-correlated with MS severity were enriched for proteins in the platelet alpha granule lumen (q-value = 5.94 e-9), fibrinogen complex (q-value = 5.03 e-5), but also endoplasmic reticulum (ER) lumen (q-value = 1.40 e-3) and azurophil/secretory granules of neutrophils (q-value = 3.40 e-3). Likewise, axon guidance and NOTCH signaling pathways dominated negative correlations with MS severity outcomes. However, these analyses also identified stronger immune signatures: Th17 cell differentiation, cytokine-cytokine interaction, and TGF-beta signaling. Consistent with the dominant role of CNS-related proteins in negative associations with MS severity, the Cellular Components Gene Ontology identified neuronal cell body (q-values from 1.96 e-5 to 3.07 e-2), axon (q-values from 1.23 e-5 to 3.99 e-2), dendrites (q-values from 9.50 e-3 to 3.99 e-2), and myelin sheath (q-values from 9.10 e-3 to 3.00 e-3) as negatively-associated with all MS severity models. Finally, visual review of STRING networks of negatively associated proteins identified VEGFA as a consistent hub across all three MS severity outcomes.

### Development and validation of CSF biomarker-based severity models

Encouraged by the fact that unbiased proteomic analysis of MS CSF measured processes previously seen in MS CNS tissue, we employed machine learning to aggregate CSF proteins into prognostic model(s) that can be used in clinical decisions.

We derived total of 3 models, each for primary, and both secondary MS severity outcomes, using the random forest algorithm (20) and a variable selection pipeline (21) (Figure 5A). Because these complex models can overfit the data, their validity must be assessed using the independent validation cohort that was not used in the pipeline development. We’ll present only the results from the independent validation cohort (Figure 5C).

**Figure 5:**
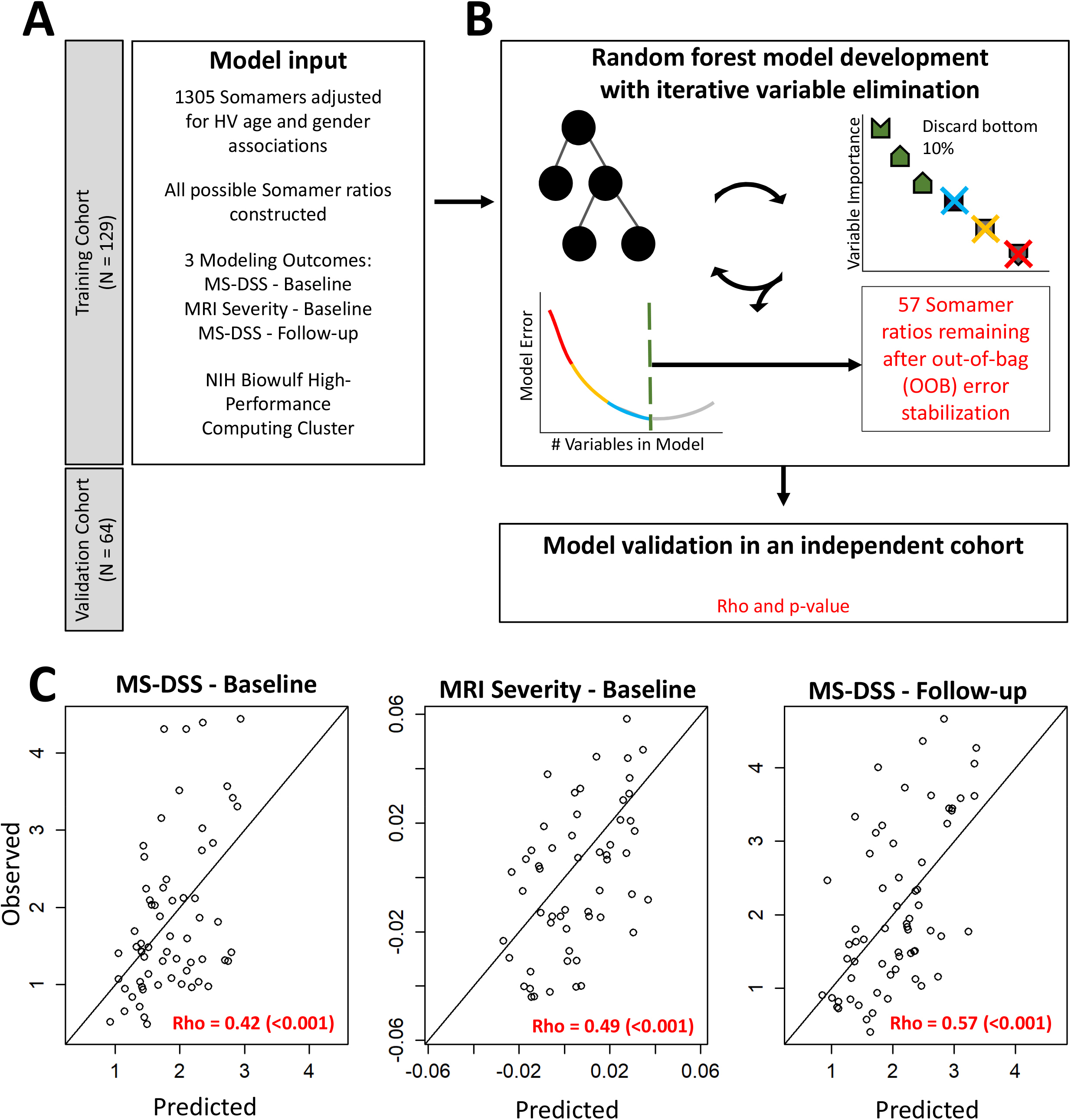
Constructing a CSF-based severity model of MS using statistical learning. **A)** Patients were split into training and validation cohorts, and all model development was performed using the training cohort. Somamers were adjusted based on healthy volunteer (HV) relationships with age and gender, and all possible Somamer ratios were constructed. Using a high-performance computing cluster, three modeling outcomes were used: multiple sclerosis disease severity score (MS-DSS) at baseline, magnetic resonance imagine (MRI) severity at baseline, and MS-DSS at most recent follow-up. **B)** Graphical depiction of the statistical learning pipeline used for model construction. In order to reduce the number of considered ratios, a statistical learning pipeline using random forest was implemented. At each step, 10 random forest models were constructed, and variable importance measures based on node impurity were averaged together. The 10% least contributing variables were excluded, and the process iterated, with the training out-of-bag (OOB) error recorded at each step. The final model was selected when the OOB error had minimized and stabilized (57 ratios for the primary outcome). **C)** Scatterplots displaying the observed severity endpoint (y-axis) versus the model predictions (x-axis) for the validation cohort. Spearman correlation coefficients and p-values are displayed in the bottom-right corner of each panel. Concordance line (x=y) is shown in black. See also Table S7.

For the primary outcome (MS-DSS baseline), the model selected 57 Somamer ratios (75 unique Somamers) and validated its predictive power in the independent cohort (rho = 0.420, p-value < 0. 001). The MRI-based MS severity model achieved slightly stronger validation (rho = 0.494, p-value < 0.001) with 21 ratios (35 unique Somamers). Finally, 21 ratios (34 unique Somamers) selected by MS severity model based on follow-up clinical data (MS-DSS Follow-up) validated the best predictive performance (rho = 0.570, p-value < 0.001).

Supplementary information contains annotated workbook (Table S7) that includes variable importance metrics (22), Spearman correlation coefficients and FDR adjusted p-values for correlations with all MS severity outcomes.

Because most research groups use EDSS-based MS severity outcomes, we assessed correlation with these MS severity outcomes not used for model development (i.e., MSSS and ARMSS). All 3 models achieved statistically-significant correlations with both EDSS-based outcomes of MS severity (Table 1; Spearman Rho ranging from 0.423 - 0.306; p-value ranging from 0.001-0.025).

Additionally, for MS-DSS baseline model, calculated in the vicinity of CSF collection, we could assess the true predictive value of CSF biomarker-based models against “future” clinical progression rates. These were calculated using patient-specific linear regressions as prospectively measured CombiWISE slopes (Figure 4A and Table 1). The reader is advised that for MS-DSS follow-up model, we are comparing de facto *past* disability progression slopes, whereas MS-DSS is optimized for predicting *future* disability progression rates (16). The CSF-biomarker based model “MS-DSS baseline” predicted prospectively measured disability slopes in the validation cohort (Spearman Rho = 0.354, p = 0.008; Table 1).

**Table 1:**
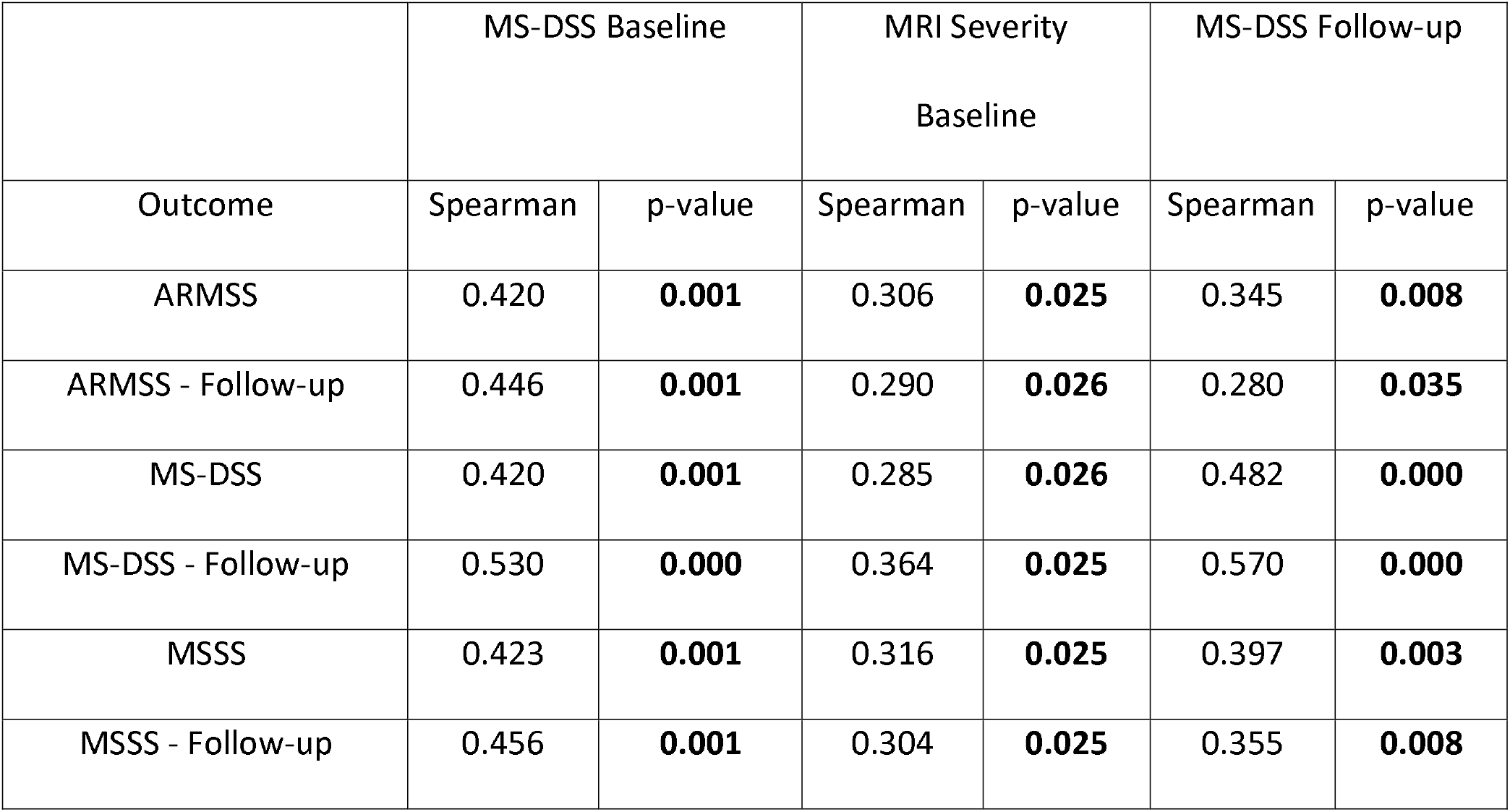
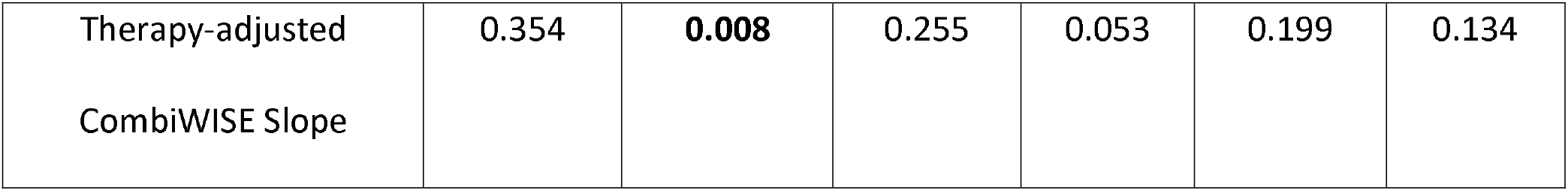
Associations between CSF model predictions and MS severity outcomes in the validation cohort. Spearman correlation coefficients and false-discovery rate (FDR) adjusted p-values between the 3 constructed CSF-based severity models (Multiple Sclerosis Disability Severity Score (MS-DSS) at baseline, MS-DSS at follow-up, and Magnetic Resonance Imaging (MRI) severity measure) across 7 MS severity outcomes: MS-DSS Follow-up, Multiple Sclerosis Severity Score (MSSS) at follow-up, Age Related Multiple Sclerosis Severity (ARMSS) at follow-up, MS-DSS at baseline, MSSS at baseline, ARMSS at baseline, and Therapy-adjusted Combinatorial weight-adjusted disability score (CombiWISE) slopes. Bold indicates an FDR adjusted p-value < 0.05. See also Table S7.

### SOMAscan-based models of MS severity reveal pathophysiological heterogeneity among MS patients that transcends clinical classification of MS subtypes

In cardiovascular diseases, different mechanisms contribute to development of disability in different patients. It is likely that such heterogeneity of disease mechanisms exist in all polygenic diseases, including MS.

To examine whether heterogeneity of pathogenic mechanisms exist among different MS patients, cluster analysis (23, 24) was performed using all Somamers selected from the three MS severity models using the ComplexHeatmap and dynamicTreeCut R packages (25) (26). Seven distinct patient clusters (Figure 6) were observed, with major differences across four protein modules: 1. Myeloid lineage/TNF module (Module 1; red annotation; Table S8), 2. CNS repair module (Module 2; green annotation; Table S9), 3. Complement/coagulation module (Module 3; blue annotation; Table S10), and 4. Adaptive immunity and CNS stress module (Module 4; black annotation; Table Sll). The protein module names were based on STRING annotations (Tables S8-11).

**Figure 6:**
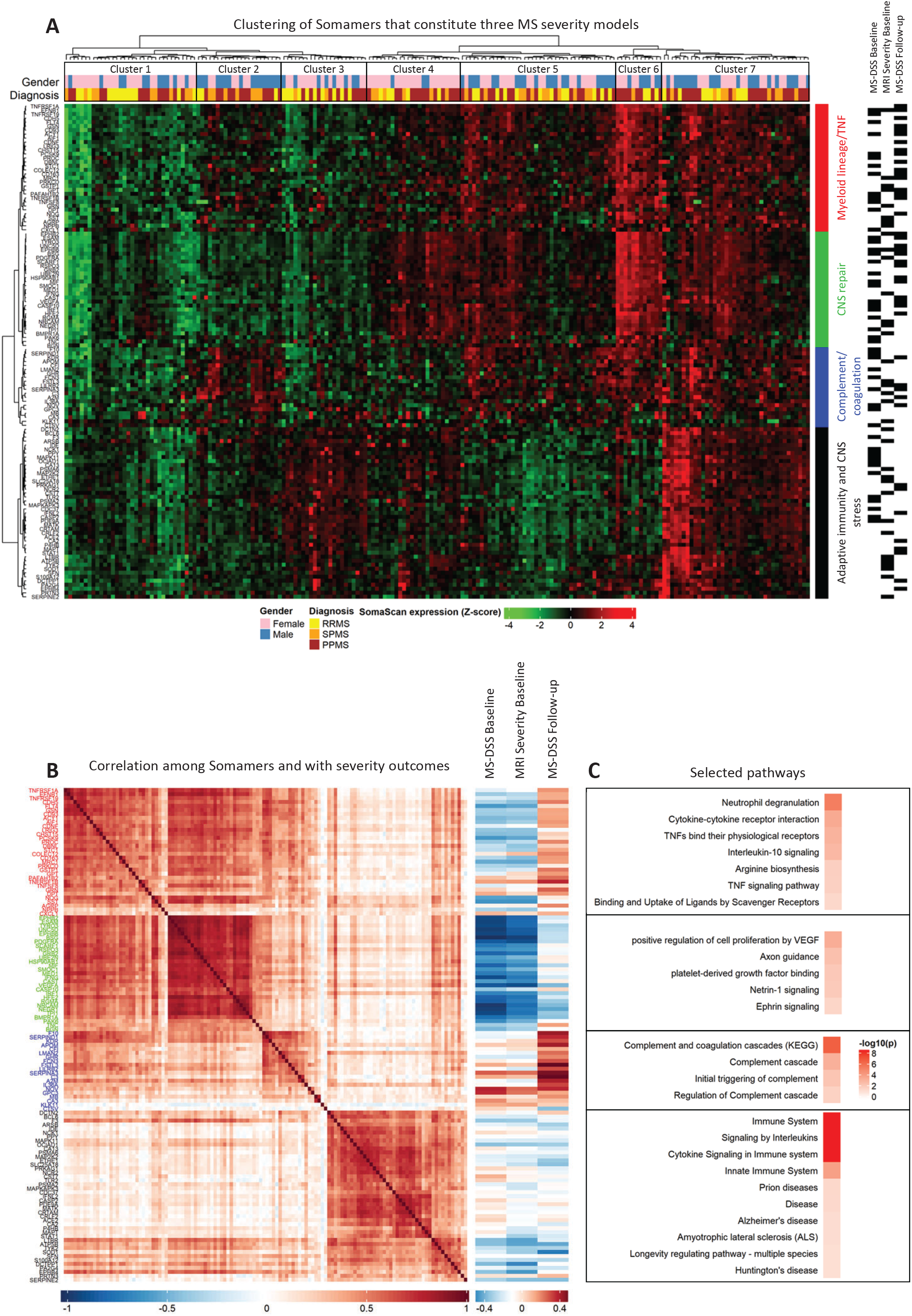
SOMAscan-based models of MS severity reveal pathophysiological heterogeneity among MS patients that transcends simple clinical classification of MS subtypes. **A)** Heatmap displaying the log-expression of the selected proteins from the 3 severity pipelines, with hierarchical cluster analysis identifying 4 protein modules (rows) across 7 patient clusters (columns). The seven patient clusters are distinguished with black boxes at the top of the heatmap. Annotations of diagnosis (yellow = relapsing remitting multiple sclerosis, orange = secondary progressive multiple sclerosis, brown = primary progressive multiple sclerosis), and gender (pink = female, blue = male) are displayed at the top of the heatmap as tiles, and Somamer module membership (red = myeloid lineage/TNF, green = central nervous system repair, blue = complement/coagulation, black = adaptive immunity and central nervous system stress) is displayed as tiles on the right side of the heatmap. Black rectangles on the right of the module annotations indicate whether the specific protein was present in a given model. **B)** Spearman correlation plot of pipeline-selected Somamers, ordered by module membership (left) along with Spearman correlation coefficients between pipeline-selected Somamers and three MS severity outcomes (right). Colors of the protein labels correspond to module membership in **A. C** Selected pathways identified using STRING analysis along with FDR-adjusted −loglO(p-values) for the four protein modules, respectively. See also Table S7, Table S8, Table S9, Table S10, and Table Sll.

All three MS severity models selected biomarkers from all 4 modules (Figure 6). While the MS clinical subtypes (i.e., RRMS, SPMS, and PPMS) were distributed across all seven patient molecular groups, patient cluster 2 had a predominance of male progressive MS patients. This cluster had relatively low expression in the CNS repair module and high expression in the Myeloid lineage/TNF module and Complement/coagulation module. Consequently, these patients had higher MS severity. In contrast, patient clusters 3 and 4 were relatively enriched for female patients. Patient cluster 3 had only high expression of protein module 4 (Adaptive immunity and CNS stress) and was enriched in RRMS subjects. Patient cluster 4 had relatively high expression of all protein modules except module 3 (Complement/coagulation module), which meant that these patients had relatively low MS severity.

## Discussion

Contemporary technological advances allow for precise measurements of thousands of genes and proteins: e.g., excellent technical reproducibility of Somascan and intra-individual stability of CSF biomarkers measured by Somascan in the absence of CNS disease was previously documented (2, 4). Enhancements in computing power allows for analyses of such omics data with complex machine learning algorithms. Yet, a recent conference on digital health (http://www.kevstonesymposia.org/index.cfm?e=web.Meeting.Program&meetingid=1654) concluded that the digital revolution failed to improve clinical outcomes thus far, identifying inaccurate clinical measures as crucial limitation for development of meaningful digital health applications. We have put vast efforts into development of granular, accurate clinical and imaging outcomes for MS using data-driven machine learning approaches guided by domain expertise (16,17, 27, 28). The current work would not be feasible without access to such an exclusive, deeply phenotyped longitudinal patient cohort. The fact that more than 10-fold higher numbers of CSF biomarkers correlated with MS-DSS than with the broadly used MSSS and ARMSS MS severity outcomes reflect gains from these outcome-development efforts. Using dissimilar (i.e., clinical and brain MRI volumetric) MS severity outcomes for CSF biomarker modeling and observing reassuringly overlapping biology strengthens the presented conclusion. Lastly, we demonstrated that the CSF biomarker model predicts *future, measured* rates of clinical disability progression in the validation cohort. Although the proportion of variance explained by this CSF-based MS severity model is modest, one must keep in mind that this CSF-biomarker-based model greatly exceeds predictive ability of broadly used clinical (i.e., MSSS and ARMSS) or genetic predictors (29). In other words, analyzing CSF in the untreated stage using Somascan predicts future rates of MS disability progression without any addition information, such as pre-existing disability measured by neurological examination or CNS tissue damage measured by MRI.

Yet, the current study also has limitations: 1. The sample sizes of the training and validation cohorts may seem small if judged by conventional outcomes. Nevertheless, the small p-values adjusted for multiple comparisons in the independent validation cohort and the reproducibility of all sensitivity analyses demonstrate that, thanks to deep phenotyping, these longitudinal cohorts reproducibly identified biological modifiers of MS severity. 2. Both training and validation cohorts were collected by the same investigators, practicing in the NIH research environment conducive to such a unique, detailed longitudinal phenotyping of patients. To prove general applicability of Somascan-based CSF biomarker models, we have established a multicenter Spinal fluid consortium for MS (SPINCOMS), which was recently funded to perform an independent validation study in the real-world academic clinical practice. This validation study should be concluded in 3 years.

Notwithstanding these limitations, the current study provides novel insights into MS pathophysiology: First, MS is not associated with accelerated physiological aging on a molecular level. In fact, age-discordant CSF proteins (i.e., decreased in healthy aging and increased in MS aging; Figure 2A) point towards re-expression of CNS-developmental pathways related to axon guidance, Netrin-1, NOTCH1, NOTCH3, and Hedgehog signaling, which likely participate in CNS repair mechanisms, as these proteins are enriched in module 2 and negatively correlate with MS severity. Additional age-discordant proteins that are increasing with MS duration are IL27RA and IL10RA; expressed in MS lesions and implicated in immunoregulation of an animal model of MS, Experimental Autoimmune Encephalomyelitis (EAE) (30, 31). This explains why these proteins are increasing in MS and why, consequently, a CSF biomarker-derived model of physiological aging underestimates chronological age in (older) patients with progressive MS. Second, our results inform on the long-standing question whether CNS tissue damage outside of MS relapses and especially at the progressive stage of MS is caused by compartmentalized inflammation or neurodegenerative mechanisms: CSF biomarkers associate MS severity with both CNS- and immune-related pathways. This study demonstrates substantial disease heterogeneity in the 4 major disease mechanism pathways represented by Somascan protein clusters among MS patients. This molecular heterogeneity of putative pathogenic mechanisms cannot be gleaned from clinical phenotypes and must be measured by CNS biomarkers. Furthermore, inflammatory mechanisms linked to MS severity do not include only adaptive immunity, targeted by most FDA-approved therapies. This study strongly links innate immunity (i.e., complement/coagulation cascade and myeloid lineage) to the rate of accumulation of disability in MS. The complement and coagulation pathways (protein module 3) were positively associated with MS severity, while developmental CNS processes associated with NOTCH signaling/myelin-related proteins (protein module 2) were negatively associated with MS severity in all three models. These results are consistent with recently validated genetic associations of complement pathway and CNS processes with MS severity (32, 33).

In contrast, proteins associated with myeloid lineage and TNF signaling (protein module 1) and those linked to adaptive immunity and response to stress (protein module 4) do not have such polarized association with MS severity: they are more diverse, with generally weaker and directionally-opposite correlations with MS severity. We interpret this as evidence that myeloid lineage and adaptive immunity does not play a uniformly detrimental role in CNS tissue destruction in MS. This interpretation is supported by experimental models, where myeloid lineage and cytotoxic lymphocytes may mediate CNS tissue destruction under certain conditions but are also essential for immunoregulation and CNS repair, including remyelination (34). This phenomenon was previously described as “beneficial autoimmunity”. For example, rapid phagocytosis of myelin debris is a pre-requisite for effective remyelination (35, 36), while autoreactive T cells mediate recovery from CNS injury, including remyelination (37). MS biopsy data support the beneficial autoimmunity concept by identifying a positive association between remyelination and the presence of T cells in early MS lesions (38).

Although it is intuitive to interpret positive associations with MS severity as detrimental and negative as beneficial, we must remember that correlation does not necessarily imply causation and that an increase in protein concentration in biological fluids results from either its increased secretion or decreased consumption, or, the combination of both. Another interpretation-limiting factor is a biased knowledge base in public databases towards cancer biology with underrepresentation of CNS processes, especially those related to neuroinflammation. For example, increased CSF levels of early complement proteins may not reflect their blood origin, but rather a proinflammatory, toxic response of microglia and astrocytes (11, 39), even though this biology was not annotated in pathway analyses. Hence, mechanistic studies must follow our results to unequivocally identify cellular sources of biomarkers assembled in Somascan predictive models and the conditions under which they are released and consumed during physiological and pathogenic interactions between CNS and immune cells.

Lastly, intra-individual heterogeneity in pathways linked to MS severity observed in this study is highly reminiscent of pathological heterogeneity involved in the formation of acute MS lesions (38). This information is essential for development of new, process-specific treatments aiming to slow CNS tissue destruction in patients with residual progression on immunomodulatory drugs. For example, activation of complement and coagulation cascade is lacking in patients belonging to clusters 1, 3, 4, activation of myeloid lineage is absent (or low) in patient-clusters 1 and 2, while biomarkers of adaptive immunity and CNS stress are underrepresented in patient-clusters 1, 2, and 5 (Figure 6). Thus, without CSF biomarker guidance, almost half of the participants in clinical trials of novel treatments may lack the target of the tested medication. This will inevitably dilute therapeutic response on a group level, requiring prohibitively large Phase 2/3 trials. Furthermore, even if such an expensive drug development succeeds, the blind application of such treatments will incur high societal cost and unnecessarily expose patients who lack therapeutic targets to the side-effects of applied drugs. Because the 2017 modification of MS diagnostic criteria include possibility of examination of CSF for OCB and because examination of CSF biomarkers is gaining popularity in other CNS diseases (40), implementation of contemporary CSF biomarker assays such as Somascan into clinical practice has a potential to revolutionize drug development and personalize treatments in patients with all neurodegenerative disorders.

## Methods

### Subjects

MS patients and HVs were prospectively recruited under an approved IRB protocol “Comprehensive Multimodal Analysis of Neuroimmunological Diseases of the Central Nervous System” (Clinicaltrials.gov identifier NCT00794352) and signed written informed consent. To be considered for the study, patients must have had a clinically definite MS diagnosis, a lumbar puncture (LP) within one year of a clinical visit that included four clinical scales (i.e., Expanded Disability Status Scale (EDSS) (41), Scripps Neurological Rating Scale (SNRS) (42), Nine-Hole Peg Test (9HPT), and Timed 25-Foot Walk (25FW)) required for calculation of Combinatorial Weight-adjusted Disability Scale (CombiWISE) (28). To assure that CSF biomarkers, imaging, and clinical data were not influenced by treatments or MS exacerbations, patients were excluded if they were in MS exacerbation or have been on low-efficacy therapies (i.e., Copaxone, interferon-beta preparations, and oral disease-modifying treatments [DMTs]) for within 3 months of LP, or high-efficacy therapies (i.e., Natalizumab, Daclizumab, Alemtuzumab, Rituximab, or Ocrelizumab) within 6 months of LP [note that the classification of drugs into low and high efficacy was adopted from a meta-analysis of age-adjusted efficacies from controlled clinical trials (43)]. HV inclusion criteria were ages 18-75, lack of neurological diagnosis or systemic disease that could influence neurological disability or brain MRI and with vital signs in the normal range during the initial screening. The demographic data of all subjects are detailed in Table S1.

### Clinical data

Patients underwent neurological examination by an MS-trained clinician. Before 2017, the calculation of neurological rating scales EDSS and SNRS was performed by each clinician. After 11/2017, the calculation of all neurological rating scales was fully automated using Digitalized Neurological examination App NeurEx on iPad (17), which also computes the NeurEx score, a continuous disability score ranging from zero to theoretical maximum of 1349. For clinical visits linked to CSF collection before 11/2017, an MS-trained clinician retrospectively transcribed the neurological examination documented in NIH electronic medical records into NeurEx App. Clinicians rating neurological disability were blinded to volumetric MRI data and CSF biomarker data as well as to calculated MS severity scales (described below).

Non-clinical investigators, blinded to neurological disability scales, MRI volumetric data, and CSF biomarker data collected timed 25 foot walk (25FW) and 9-hole peg test (9HPT) and uploaded these to the research database. All clinical and functional data were quality controlled during weekly clinical care meetings after which the corresponding parts of the database were locked for modification.

CombiWISE was automatically computed in the research database from EDSS, SNRS, 25FW and 9HPT values as described (28). Machine learning-optimized MS Disease Severity Scale (MS-DSS) was computed as described Weideman, et al. (16). MS-DSS has higher accuracy in predicting future rates of disability progression in comparison to EDSS-based severity scales - MS Severity Score (MSSS (14)) and Age Related MS Severity Score (ARMSS (15)).

All computed scales developed by the Bielekova lab are freely available at https://bielekovalab.shinyapps.io/msdss/. NeurEx software is likewise freely available to non-commercial entities.

### CSF processing

CSF was collected on ice and processed according to a written standard operating procedure by investigators blinded to clinical and MRI outcomes. Aliquots were assigned alphanumeric identifiers and centrifuged for 10 minutes at 4 °C within 15 minutes of collection. Until use supernatant was aliquoted and stored in polypropylene tubes at −80 °C.

### SOMAscan

CSF samples were analyzed blindly, using SOMAscan technology (SomaLogic, Boulder, CO), a DNA-aptamer-based assay that measures relative fluorescence units (RFUs) of 1,305 proteins (available after October 2016, referred to as the 1.3K platform) by the NIH Center for Human Immunology. In total, 193 MS patients and 24 HVs (42 unique samples) had CSF samples available on the 1.3K platform that met the inclusion criteria discussed above.

### Magnetic resonance imaging (MRI)-based MS severity scale

The brain MRIs were performed on 1.5T and 3T Signa units (General Electric, Milwaukee, WI) and 3T Skyra (Siemens, Malvern, PA) equipped with standard 16- and 32-channel imaging coils. MRI sequences used for grading comprised of T1 magnetization-prepared rapid gradient-echo (MPRAGE) or fast spoiled gradient-echo (FSPGR) and T2 weighted three-dimensional fluid attenuation inversion recovery (3D FLAIR). The details of the MRI sequences is previously published (32).

The brain MRI images were evaluated by two complementary methods: 1. semiquantitative ratings were assembled to Combinatorial MRI scale of CNS tissue destruction (COMRIS-CTD) using the published formula (32), available at https://bielekovalab.shinvapps.io/msdss/. 2. Identical MRI scans were analyzed using LesionTOADS volume segmentation algorithm (44), performed internally at the NIH until December 2018 and afterwards in collaboration with QMENTA medical image-processing platform https://www.qmenta.com/.

Raw unprocessed but locally anonymized and encrypted T1 - MPRAGE or T1 - FSPGR and T2 - 3D FLAIR DICOM files as input sequences, ideally with 1 mm^3^ isotropic resolution, were uploaded to the QMENTA platform. LesionTOADS, now implemented into the cloud-based service, is a fully automated segmentation algorithm using multichannel MRI data (45). The uploaded sequences are anterior commissure-posterior commissure (ACPC) aligned, rigidly registered to each other and skull stripped (the T1 image is additionally bias-field corrected). The segmentation is performed by using an atlas-based technique combining a topological and statistical atlas resulting in computed volumes for each segmented tissue in mm^3^.

To calculate the MRI severity measurement, brain atrophy measured as 1-brain parenchymal fraction (BPFr, calculated as proportion of intracranial volume occupied by brain tissue; [Cortical gray matter + Caudate + Thalamus + Putamen + Normal appearing white matter + Lesions)/(Cerebrum gray matter + Caudate + Thalamus + Putamen + Normal appearing white matter + Lesions + Sulcal CSF + Ventricular CSF]) was regressed against age using baseline data in the full cohort of patients with MS. This demonstrated strong evidence of increasing brain atrophy over increasing age (t_12_s = 4.41, p-value=0.00002). The residuals from the resulting regression were then calculated. These residuals were used as the MRI severity measure, where positive values are indicative of faster CNS tissue destruction in a manner analogous to clinical measures of MS severity.

### Adjusting Somamers for differences in age and gender

As previous studies (1, 46, 47) have demonstrated associations between specific CSF proteins measured by SOMAscan and confounding factors age and gender in HVs, we sought to adjust protein levels in our MS patients to account for natural physiological differences due to age and gender. An initial list of Somamers were selected from published INTERVAL cohort examining serum proteins using Somascan (1) where either age or gender associations were detected. The natural log of these Somamers were modeled using regression to test for age and/or gender difference in the 1.3K platform in CSF samples from MS patients as well as HVs. Somamers with an association between age and/or gender with p<0.05 in the MS cohort and concordant directions between INTERVAL HV serum and HV CSF were adjusted in the MS data using regression models derived from HV CSF samples.

### Examining individual associations between Somamers and disease outcomes

Individual Spearman correlations were computed between adjusted protein levels and MS severity endpoints. All p-values for individual Somamer correlations were adjusted for multiple comparisons using the FDR method (18). See also Table S6.

### Constructing a CSF-based severity model of MS using statistical learning

Random forest models (20) using the ranger R package (48, 49) were used to construct the CSF-biomarker-based models of MS severity. For each platform, the CSF samples at the untreated baseline were used to predict MS-DSS (see clinical data Methods section), at both the baseline visit and the most recent follow-up, and the MRI severity measure at baseline. All possible protein ratios were included in the modeling along with individual markers. The (4) explains the principle of Random Forests algorithm and rationale for using protein ratios.

Prior to model development, the available data were randomly split into training and validation cohorts, with 2/3 being used as a training cohort (N=129) and 1/3 being retained only for model validation (N=64). To reduce number of ratios/markers based on predictive performance, a variation of the published procedure Calle, et al. (21) was performed (Figure 5B). Briefly, 10 random forests were run using the training cohort and variable importance measures based on node impurity (22) were averaged together. The bottom 10% of variables according to these average variable importance measures were removed from the candidate set. This process was repeated until only 3 variables remained. The mean and standard deviation of the out-of-bag (OOB) error was graphically assessed to determine the final cut point for each model. This procedure was performed for Somamers adjusted for age and gender. For each instance, a final random forest model was constructed in the training cohort. Biological interpretations of the selected proteins were explored using cluster analysis (23, 24) and STRING analysis (19).

## Statistics

All statistical tests are two-sided. All correlations were calculated using Spearman correlation coefficients. Therefore, no assessment of linearity was performed. When determining if Somamers had physiological age and gender associations, t-statistics from multiple linear regression models were constructed. When comparing differences between CSF predicted age and observed age between diagnosis, ANOVA followed by all pairwise comparisons using Tukey’s honest significant difference. ANOVA assumptions were assessed using residual versus fitted value plots and were satisfied.

## Study approval

All subjects were prospectively recruited under an approved IRB protocol “Comprehensive Multimodal Analysis of Neuroimmunological Diseases of the Central Nervous System” (Clinicaltrials.gov identifier NCT00794352) and signed written informed consent.

## Data Availability

Raw data will be available upon publishing of the peer-reviewed manuscript.

## Author contributions

Study concept and design: B.B.; data acquisition and analysis: B.B., C.B. P.K. M.V, M.G.; drafting of the manuscript and figures: all authors.

## Acknowledgments

This study was funded by the Intramural Research Program of the National Institute Allergy and Infectious Diseases (NIAID) of the National Institutes of Health (NIH). This work utilized the computational resources of the NIH HPC Biowulf cluster, (http://hpc.nih.gov).

## Supplemental information

**Table S1:** *Demographic data for the training and validation cohorts in addition to control data. P-value (not adjusted for multiple testing) column tests for difference in specific demographic parameter with respect to the two cohorts (excluding controls). For gender, a chi-squared test of independence was used. For quantitative variables, a Wilcoxon rank test was used. Note although there were 24 healthy volunteers, some had more than one unique sample. There was 42 unique healthy volunteer samples. HV = healthy volunteer, RRMS = relapsing-remitting multiple sclerosis, SPMS = secondary progressive multiple sclerosis, PPMS = primary progressive multiple sclerosis, EDSS = expanded disability status scale, MS-DSS = multiple sclerosis disability severity score. See also methods section*.

**Table S2:** *Search Tool for the Retrieval of Interacting Genes/Proteins (STRING) annotation for age associated Somamers with concordant directionality between healthy volunteers and multiple sclerosis patients. See also Figure 1*.

**Table S3:** *Search Tool for the Retrieval of Interacting Genes/Proteins (STRING) annotation for age associated Somamers with discordant directionality between healthy volunteers and multiple sclerosis patients. See also Figure 1*.

**Table S4:** *Search Tool for the Retrieval of Interacting Genes/Proteins (STRING) annotation for Somamers that are elected in females. See also Figure 2*.

**Table S5:** *Search Tool for the Retrieval of Interacting Genes/Proteins (STRING) annotation for Somamers that are elected in males. See also Figure 2*.

**Table S6:** *Spearman correlations and False Discovery Rate adjusted p-values between all Somamers and various multiple sclerosis severity measures in untreated patients. See also STAR methods*.

**Table S7:** *Annotated Somamer ratios for three statistical learning models of multiple sclerosis severity. See also Figure 5, Figure 6 and Table 1*.

**Table S8:** *Search Tool for the Retrieval of Interacting Genes/Proteins (STRING) annotation for Somamers present in the myeloid lineage/TNF module or “module 1”. See also Figure 6*.

**Table S9:** *Search Tool for the Retrieval of Interacting Genes/Proteins (STRING) annotation for Somamers present in the central nervous system (CNS) repair module or “module 2”. See also Figure 6*.

**Table S10:** *Search Tool for the Retrieval of Interacting Genes/Proteins (STRING) annotation for Somamers present in the complement/coagulation module or “module 3”. See also Figure 6*.

**Table S11:** *Search Tool for the Retrieval of Interacting Genes/Proteins (STRING) annotation for Somamers present in the adaptive immunity and central nervous system (CNS) stress module or “module 4”. See also Figure 6*.

